# Pseudo-Likelihood Based Logistic Regression for Estimating COVID-19 Infection and Case Fatality Rates by Gender, Race, and Age in California

**DOI:** 10.1101/2020.06.29.20141978

**Authors:** Di Xiong, Lu Zhang, Gregory L. Watson, Phillip Sundin, Teresa Bufford, Joseph A. Zoller, John Shamshoian, Marc A. Suchard, Christina M. Ramirez

## Abstract

In emerging epidemics, early estimates of key epidemiological characteristics of the disease are critical for guiding public policy. In particular, identifying high risk population subgroups aids policymakers and health officials in combatting the epidemic. This has been challenging during the coronavirus disease 2019 (COVID-19) pandemic, because governmental agencies typically release aggregate COVID-19 data as marginal summary statistics of patient demographics. These data may identify disparities in COVID-19 outcomes between broad population subgroups, but do not provide comparisons between more granular population subgroups defined by combinations of multiple demographics.

We introduce a method that overcomes the limitations of aggregated summary statistics and yields estimates of COVID-19 infection and case fatality rates — key quantities for guiding public policy related to the control and prevention of COVID-19 — for population subgroups across combinations of demographic characteristics. Our approach uses pseudo-likelihood based logistic regression to combine aggregate COVID-19 case and fatality data with population-level demographic survey data to estimate infection and case fatality rates for population subgroups across combinations of demographic characteristics.

We illustrate our method on California COVID-19 data to estimate test-based infection and case fatality rates for population subgroups defined by gender, age, and race and ethnicity. Our analysis indicates that in California, males have higher test-based infection rates and test-based case fatality rates across age and race/ethnicity groups, with the gender gap widening with increasing age. Although elderly infected with COVID-19 are at an elevated risk of mortality, the test-based infection rates do not increase monotonically with age. LatinX and African Americans have higher test-based infection rates than other race/ethnicity groups. The subgroups with the highest 5 test-based case fatality rates are African American male, Multi-race male, Asian male, African American female, and American Indian or Alaska Native male, indicating that African Americans are an especially vulnerable California subpopulation.

## 1. Introduction

Severe acute respiratory syndrome coronavirus 2 (SARS-CoV-2) has spread from its zoonotic origins in Hubei Province, China, causing a global pandemic of coronavirus disease 2019 (COVID-19) [1, 2]. As of June 12, 2020, COVID-19 has infected over 7 million people across 188 countries and regions [3]. In the early stages of an emerging epidemic such as COVID-19, estimating the infection rate (IR) and case fatality rate (CFR) of the infectious disease is of utmost importance to health officials, policy makers, and the population at large. Accurate population and subgroup estimates of CFRs provide an evidence-based rationale for policies designed to mitigate the spread of the infectious disease, help identify disparities in disease vulnerability, and inform resource allocation to communities in greatest need.

Official COVID-19 data released by governmental health agencies and other public sources are prohibited by U.S. law from containing personally identifiable information. Consequently, these data are generally summarized in an aggregate format that comprise only marginal or limited bivariate summary statistics of patient demographics, providing valuable but limited information on the heterogeneity of patient attributes. Indeed, in New York City, the epicenter of the COVID-19 outbreak in the U.S., the reported infection rates and case fatality rates for African Americans were disproportionately higher than other races, according to data released by the New York City Department of Health and Mental Hygiene [4]. Data from several other U.S. states, including New Jersey[5], California[6], and Illinois[7], exhibited similar trends. Gender and age-disaggregated national case data from a vast array of countries across the globe reveal that males and older individuals generally have substantially higher case fatality rates. Furthermore, evidence from numerous clinical studies of COVID-19 risk factors have established that gender and age are risk factors for COVID-19 infection mortality [8, 9, 10, 11]. However, by aggregating, data from governmental health agencies or other public sources do not provide granular information on the combined effect of the risk factors under consideration. In particular, how IRs and CFRs vary across population subgroups characterized by gender, age, and race jointly has not yet received substantial attention. Understanding the gender-age-race dynamics of COVID-19 infection and mortality would provide deeper insights into the disparities that exist in the effects of COVID-19 on the population.

Various methods for using information contained in aggregate data have been proposed in a wide array of applications [see, e.g. 12, 13, 14], and there is growing interest in leveraging marginal summary statistics in publicly released COVID-19 datasets to quantify the impact of various risk factors on COVID-19 mortality [15]. In this paper, we propose a method that helps overcome the limitations of having only aggregate summary statistics on COVID-19 cases and fatalities to obtain early estimates of COVID-19 IRs and CFRs for population subgroups defined by combinations of risk factors.A major difference between the prevalent approaches to analyzing marginalized data and our proposed method is that we incorporate multivariate population demographic data, which provides estimates of the joint probability distribution of risk factors for the disease. Specifically, we propose a pseudo-likelihood based multivariable logistic regression approach that combines publicly released aggregate COVID-19 case and fatality data with multivariate population-level demographic survey data.

The proposed method is composed of two main steps. First, we model COVID-19 IRs using a multivariable logistic regression model, estimating its parameter values from publicly available COVID-19 case data. Second, we estimate COVID-19 CFRs based on the recovered IRs and publicly available COVID-19 fatality data. This paper uses California as an example case study, but the approach is easily generalized to other states. We carry out the analysis using the most recent COVID-19 case and fatality data from the California Department of Public Health (CDPH) [6, 16, 17] and population-level demographic data from the California Health Interview Survey (CHIS) [18] to obtain estimates of IRs and CFRs for subgroups of the California state population characterized by the joint distribution of gender, age, and race. Not every person who may be infected with COVID-19 is tested, and in some locations only people who are symptomatic are tested. This introduces sampling bias into the COVID-19 data that prevents straightforward estimation of the true IRs and CFRs. To circumvent this issue, we estimate test-based IR (T-IRs) and test-based CFRs (T-CFRs) that depend on the availability and use of testing and may differ from the true IRs and CFRs. In particular, we expect true IRs to be greater than T-IRs and true CFRs to be less than T-CFRs due to the presence of asymptomatic and undiagnosed infections. While the test-based rates do not estimate the overall population rates, they capture the vast majority of severe or fatal COVID-19 infections, because these individuals are very likely to be tested. Consequently, the T-IRs and T-CFRs estimated by our method provide valuable insights into the disparities in COVID-19 outcomes that exist across gender, age, and race/ethnicity groups and furnish guidance for public policy related to the control and prevention of COVID-19.

## 2. Data

Our method for estimating COVID-19 T-IRs and T-CFRs relies on two data sources: daily COVID-19 data for California from the California Department of Public Health (CDPH), and the 2017–2018 wave of the California Health Interview Survey (CHIS).

CDPH data are publicly available and provide up-to-date information on the number of COVID-19 cases and fatalities in California by gender [16], age [17], and race/ethnicity [6], separately. The case and fatality data as of June 12, 2020 are presented in Table 1. CDPH divides the population into ten age groups: less than 5, 5–17, 18–34, 35–49, 50–59, 60–64, 65–69, 70–74, 75–79, and 80 and above. Age group is missing from less than 0.1% of the confirmed cases and deaths reported by CDPH. The eight race and ethnicity groups in the publicly released dataset are LatinX/ Hispanic (LatinX), White/ Caucasian (White), Asian, African American/Black (AA), Multi-Race, American Indian or Alaska Native (AIAN), Native Hawaiian and other Pacific Islander, and others. We combined the last two race and ethnicity groups due to their small size in the California population. Race and ethnicity are missing in almost 29% of confirmed cases and 1% of the deaths. The CDPH data also provide the number of COVID-19 cases and fatalities by race and age jointly [6], the only state in the U.S. doing so at the time of writing.

**Table 1:** Confirmed COVID-19 cases and fatalities by gender, age and race/ethnicity in California as of June 13, 2020 [16, 17, 6]

| **Demographic** | **Group** | **Confirmed Cases** | **Deaths** | **Death Rate (%)** |
| --- | --- | --- | --- | --- |
| **Gender** | **Male** | 73,480 | 2,727 | 3.71% |
|  | **Female** | 71,501 | 2,146 | 3.00% |
| **Age Group** | **0–17** | 9,819 | 0 | 0.00% |
|  | **18–34** | 41,766 | 52 | 0.12% |
|  | **35–49** | 36,675 | 244 | 0.67% |
|  | **50–59** | 23,486 | 439 | 1.87% |
|  | **60–64** | 9,297 | 339 | 3.65% |
|  | **65–69** | 6,735 | 450 | 6.68% |
|  | **70–74** | 4,980 | 506 | 10.16% |
|  | **75–79** | 3,841 | 601 | 15.65% |
|  | **80+** | 8,847 | 2,247 | 25.40% |
| **Race & Ethnicity** | **LatinX** | 58,415 | 1,941 | 3.32% |
|  | **White** | 18,783 | 1,594 | 8.49% |
|  | **Asian** | 8,271 | 709 | 8.57% |
|  | **AA** | 4,906 | 458 | 9.34% |
|  | **Multi-race** | 775 | 33 | 4.26% |
|  | **AIAN** | 201 | 13 | 6.47% |
|  | **Other** | 12,389 | 58 | 0.47% |

To supplement the CDPH COVID-19 data, we used demographic data on the California population collected by the California Health Interview Survey (CHIS). CHIS is the largest state health survey in the U.S., conducted by the UCLA Center for Health Policy Research in collaboration with the California Departments of Public Health and Health Care Services. CHIS interviews over 20,000 Californians each year, collecting information on a wide range of demographic and health variables. CHIS oversamples certain population subgroups to achieve more reliable and precise estimates for these subgroups, and estimates a sampling weight for each respondent to represent the reciprocal of the probability of selection. We use the 2017–2018 wave of CHIS in our analysis, which consists of 45,369 subjects interviewed, focusing on the following three demographic variables recorded: gender, age, and race and ethnicity groups.

## 3. Methods

### 3.1. Infection Rate Estimation Procedure

We propose estimating COVID-19 T-IRs given gender, age and race using a multivariable logistic regression model. The variables we use in our analysis are listed in Table 2. Letting age 0–17, female, and LatinX be the reference categories, we let ***𝓏*** *∈* {0, 1}^*p*^*∈* ℝ^*p*^ denote the gender-age-race covariate setting of the covariates ***Z*** in Table 2, where *p* is 15. The postulated IR model follows

**Table 2:** Variables used in the infection rate estimation procedure

| Variable | Definition |
| --- | --- |
| Infection Status | Dichotomous outcome indicating COVID-19 infection<br>(0 = No, 1 = Yes) |
| Gender | Dichotomous covariate indicating male or female<br>(0 = Female, 1 = Male) |
| Age | Categorical covariate with the following age groups:<br>0–17 (reference level), 18–34, 35–49, 50–59, 60–64, 65–69,<br>70–74, 75–79, 80+ |
| Race/Ethnicity | Categorical covariate with the following race categories:<br>LatinX (reference level), White, Asian,<br>African American/Black, Multi-Race,<br>American Indian or Alaska Native, Other |

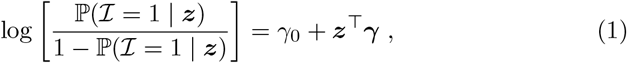

where *ℐ ∈* {0, 1} represents infection status, *γ*_0_ is the log odds of infection for the female age 0–17 Latinx group, and ***γ*** *∈* ℝ^*p*^are the log odds ratios of infection associated with the other demographic categories.

The CDPH data provide the gender, age, and race distributions of COVID-19 infections separately [16]. To estimate the T-IRs given gender, age, and race jointly, we employ a pseudo-likelihood approach that maximizes a likelihood function constructed from univariate logistic regression models obtained by marginalizing over the covariates. The proposed method begins by first expressing Equation (1) in terms of the probability of infection conditional on the covariates,

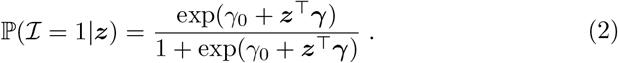

We introduce 𝕡_***X***_ (***x***) as the probability mass function of a *p*^*\**^-dimensional discrete random variable ***X*** with support 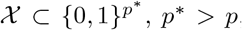, that represents the proportion of the California population with gender-age-race attributes ***x***, which is simply an augmentation of the covariate setting ***z*** in (1) to include the reference levels listed in Table 2. In other words, there exists a bijection ***𝓏*** = ***𝓏*** (***x***) from the ***X***-space to the ***Z***-space. We then define the conditional probability mass function of ***X***_*−i*_ given 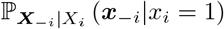, where *X*_*i*_ is the *i*^th^ element of ***X***, and ***X***_*−i*_ is the subset of ***X*** that omits *X*_*i*_. Defining 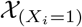 to be the subset of *𝒳* with the constraint that *X*_*i*_ = 1 and taking the expectation of both sides of Equation (2) conditional on *X*_*i*_ = 1, by the Law of Iterated Expectations we have

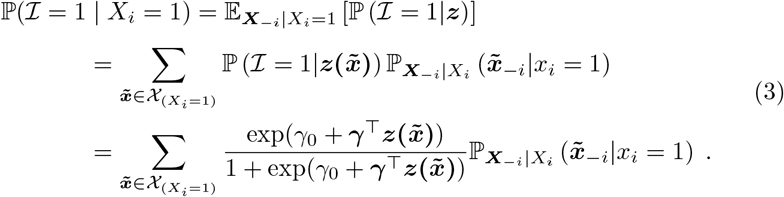

Next, we construct the individual pseudo-log-likelihoods corresponding to each univariate logistic regression of *ℐ* on *X*_*i*_ = 1 for each *X*_*i*_ *∈* ***X***. Let *N* denote the total population size, *N*_*i*1_ denote the number of individuals in the population with *X*_*i*_ = 1, and 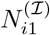 denote the total number of individuals with *X*_*i*_ = 1 who have been or will be infected with COVID-19. Therefore, 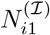 follows a binomial distribution,

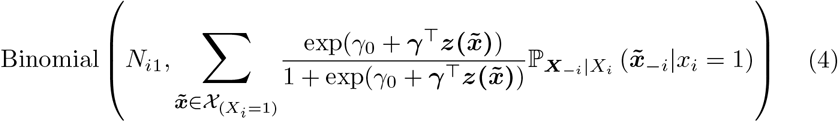

for *i* = 1, …, *p*^*\**^. We define the individual pseudo-log-likelihood of (*γ*_0_, ***γ***) for *X*_*i*_ corresponding to the binomial distribution (4) as 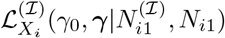, and we define the full pseudo-log-likelihood of (*γ*_0_, ***γ***) as the sum of the individual pseudo-log-likelihoods

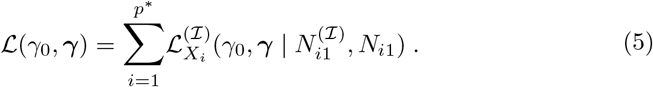

We use the CHIS data to approximate ℙ_***X***_ (***x***), which we denote 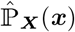. Let *N* ^(*ℐ*)^ denote the total number of individuals in the population who have been or will be infected with COVID-19, and let *π*_*ℐ*_ = 𝕡(*ℐ* = 1) denote the overall infection rate in the population. Thus, the total population size is *N* = *N* ^(*ℐ*)^*/π*_*ℐ*_. From the CDPH data presented in Table 1, we have the cumulative number of reported COVID-19 infections as of June 12, 2020, which we denote 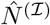. Because 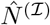 measures the cumulative number of COVID-19 infections up to June 12, 2020, and increases daily, 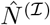 is smaller than *N* ^(*ℐ*)^, perhaps substantially. Furthermore, *π*_*ℐ*_ is unknown, and for a given estimate 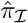 of *π*_*ℐ*_, we define 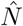 to be 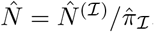. Therefore, even for accurate estimates of 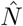 will be smaller, perhaps substantially, than the total number of individuals in the population. However, we assume here that the relative size of 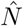 to *N* is approximately equal to the relative size of 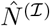 to *N*^(*ℐ*)^. Hence, 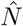 may be interpreted as an appropriately scaled version of *N* with respect to 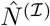 and 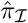 as of June 12, 2020. Likewise, we define 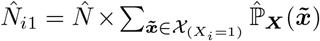 with 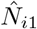 having the same interpretation as 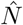 but for the subset of the population with *X*_*i*_ = 1. We denote 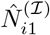 to be the cumulative number of infected individuals with *X*_*i*_ = 1 as of June 12, 2020, and present 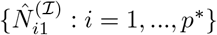 in Table 1.

Substituting 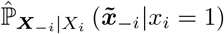 for 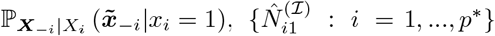 for 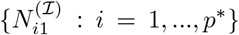 and 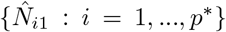 for {*N*_*i*1_ : *I =* 1, …, *p*^*∗*^} in the pseudo-log-likelihood (5), we obtain an approximate pseudo-likelihood

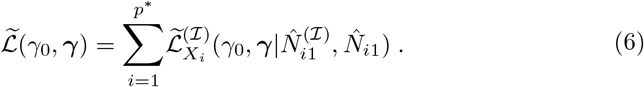

We maximize the approximate pseudo-likelihood (6) with respect to (*γ*_0_, ***γ***) to obtain our estimates

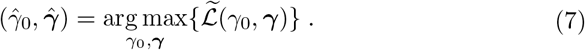

Lastly, by plugging 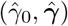 into Equation 2, we obtain the predicted test-based infection probabilities for individuals with gender-age-race covariate setting ***𝓏***

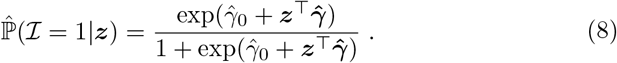

### 3.2. Case Fatality Rate Estimation Procedure

Similar to the T-IR estimation method, we model the T-CFRs given gender, age, and race using a multivariable logistic regression model. The gender-age-race covariate we use for CFR estimation (see Table 3) is the same as the covariate we use for IR estimation, except that we combined the 0–17 and 18– 34 age groups due to low numbers of fatalities among the 0–17 age group. With a slight abuse of notation, we denote ***𝓏*** *∈* {0, 1}^*q*^ *∈* ℝ^*q*^ to be the covariate setting of the vector of non-reference group covariates ***Z***, where *q* = 14. The corresponding random variable ***X*** and its covariate setting ***x*** are as defined in the preceding subsection and have dimension *q*^*\**^, where *q*^*\**^ = 17. We give the T-CFR model as

**Table 3:** Variables used in the case fatality rate estimation procedure

| Variable | Definition |
| --- | --- |
| Mortality Status | Dichotomous outcome indicating fatality status<br>(0 = No, 1 = Yes) |
| Gender | Dichotomous covariate indicating gender<br>(0 = Female, 1 = Male) |
| Age | Categorical covariate with the following age groups:<br>0–34 (reference level), 35–49, 50–59, 60–64, 65–69, 70–74,<br>75–79, 80+ |
| Race/Ethnicity | Categorical covariate with the following race categories:<br>LatinX (reference level), White, Asian,<br>African American/Black, Multi-Race,<br>American Indian or Alaska Native, Other |

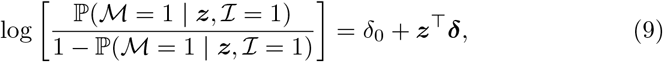

where *ℳ ∈* {0, 1} represents mortality status, *δ*_0_ is the log odds of mortality for the LatinX female age 0–34 group, and ***δ*** *∈* ℝ^*q*^ are the log odds ratios of mortality for other covariate settings.

We again employ a pseudo-likelihood approach to estimate (*δ*_0_, ***δ***) that maximizes a likelihood function constructed from univariate logistic regression models. Following similar steps as shown in the preceding subsection, we have

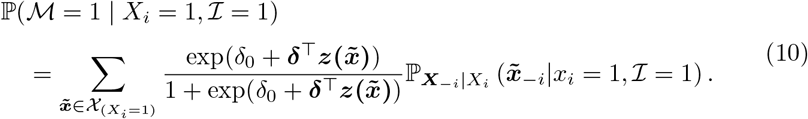

We use the CHIS data and the IR model (1) with coefficient estimates (7) to estimate 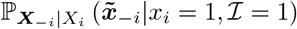. First, we estimate 𝕡(***𝓏***|*ℐ* = 1) using Bayes’ Rule

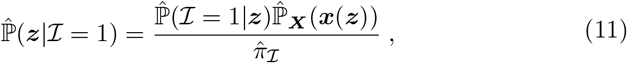

where 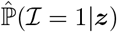 comes from Equation (8), and 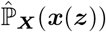 is obtained from the CHIS dataset. Then, by the definition of conditional probability, we estimate 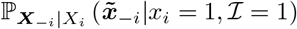 by

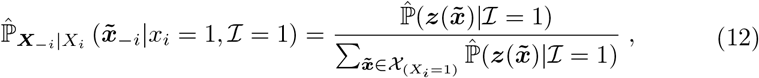

where 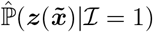 comes from Equation (11).

Analogous to the IR model, we denote 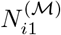 to be the number of individuals with *X*_*i*_ = 1 who have died or will die from COVID-19. Therefore, 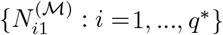 each follows a binomial distribution

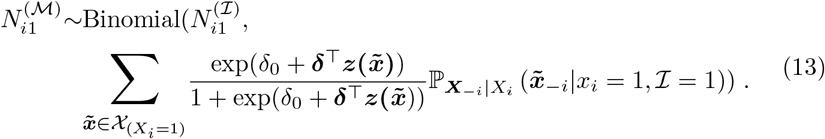

We then construct the full pseudo-log-likelihood of (*δ*_0_, ***δ***) as the sum of the individual pseudo-log-likelihoods of (*δ*_0_, ***δ***) for *X*_*i*_ corresponding to binomial distribution (13)

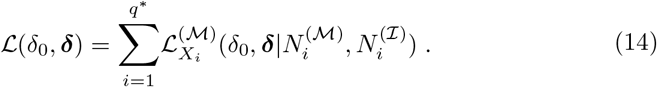

From the CDPH data presented in Table 1, we have the cumulative number of COVID-19 deaths by gender, age, and race. We denote 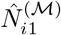 to be the cumulative number of reported deaths of infected individuals with *X*_*i*_ = 1 as of June 12, 2020. Analogous to the infection risk model, we assume that the relative size of 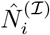 to 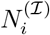 is approximately equal to the relative size of 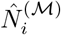 to 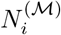. Substituting 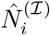 for 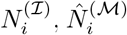 for 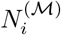, and 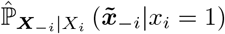 for 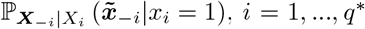, in the pseudo-likelihood (14), we obtain an approximate pseudo-likelihood

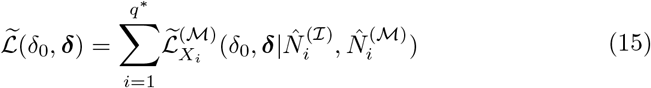

and maximize it with respect to (*δ*_0_, ***δ***) to obtain our estimates

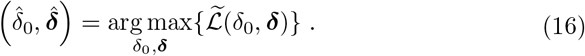

Lastly, from 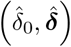, we can obtain the predicted COVID-19 test-based case fatality rates for individuals with gender-age-race covariate setting ***𝓏***,

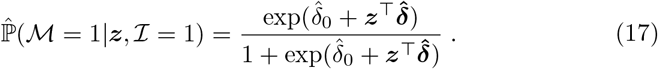

### 3.3. Monte Carlo Simulation Procedure

To quantify the uncertainty of the T-IR and T-CFR estimates in (8) and (17), respectively, we carry out a Monte Carlo procedure that repeatedly performs the T-IR and T-CFR estimation procedures described in Sections 3.1 and 3.2 sequentially, introducing sampling variation in the data in three stages. The first stage bootstraps the CHIS data with selection probabilities proportional to the sampling weights. The second stage introduces variation in 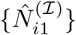 immediately prior to maximizing the approximate pseudo-log-likelihood (9), by simulating values of 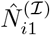 for each *i* independently from a binomial distribution with success probability equal to 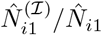, i.e.,

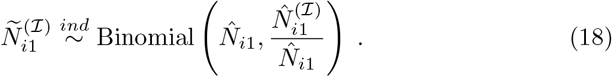

Similarly, the third stage introduces variation in the 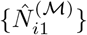 prior to maximizing the approximate pseudo-log-likelihood (15) by simulating values of 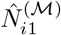 for each *i* independently from a binomial distribution with success probability equal to 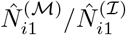,

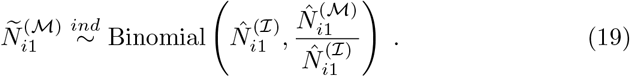

The entire Monte Carlo simulation procedure can be summarized in 5 steps:

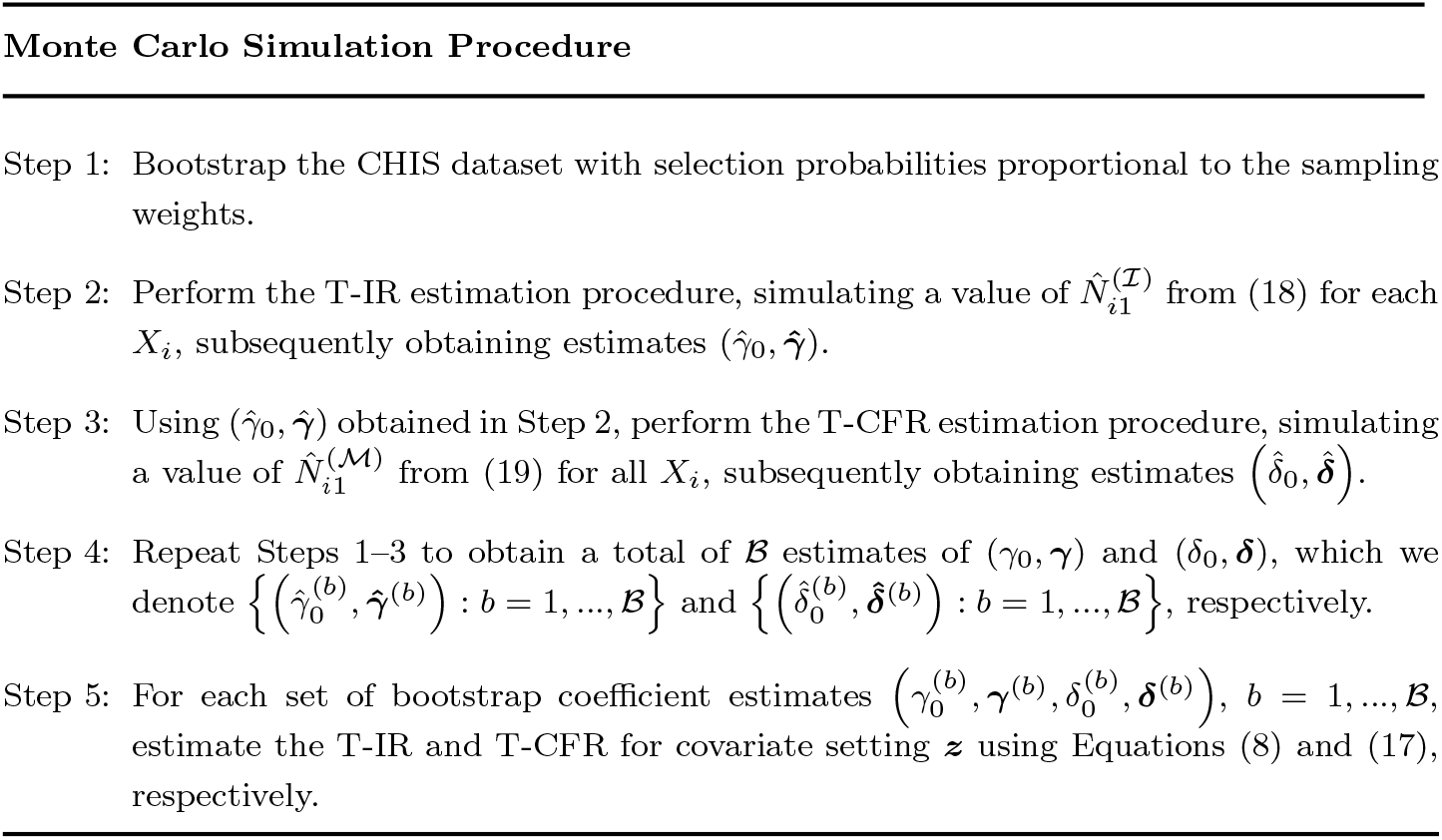

In Figure 1, We illustrate the Monte Carlo simulation procedure in a flow chart.

**Figure 1:**
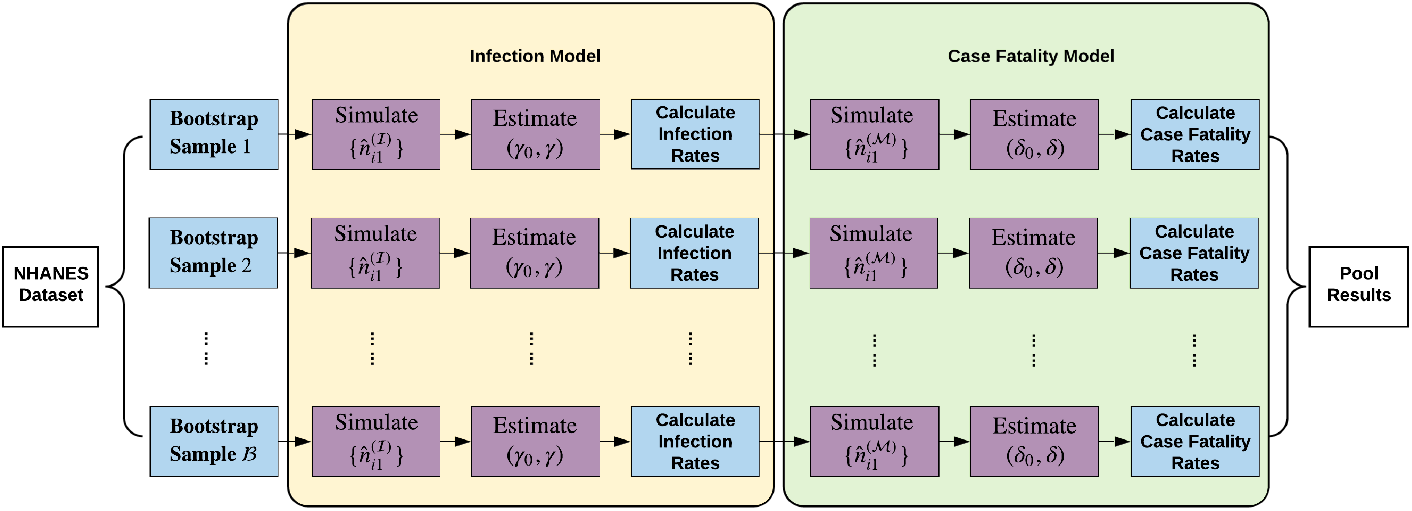
Flow chart depicting the Monte Carlo simulation procedure

### 3.4. Summary Statistics for Infection and Case Fatality Rate Estimates

In addition to estimating the T-IR and T-CFR for specific covariate settings ***𝓏*** through Equations (8) and (17), respectively, we can provide collapsed estimates of T-IRs and T-CFRs for specific values of any subset 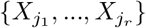 of ***X***. Let *J*_*r*_ = {*j*_1_, …, *j*_*r*_}, where *j*_*r*_ ⊂ {1, …, *p*\*};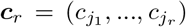, where 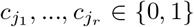; and 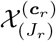 denotes the subset of *𝒳* with the constraint that 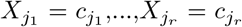. Estimates of collapsed T-IRs given 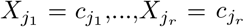 can be obtained using the marginalization formula

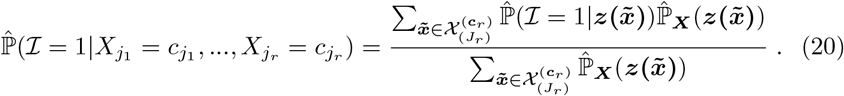

Likewise, collapsed estimates of T-CFRs given 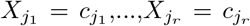, can be obtained using the marginalization formula

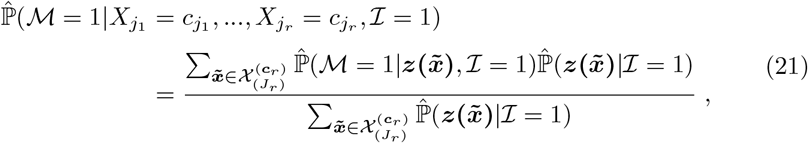

where 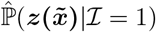 comes from Equation (11).

## 4. Results

We present select estimates of T-IRs and T-CFRs obtained from our IR (1) and CFR (8) models fit to the California data described in Section 2 and summarized according to Section 3.4. All standard errors were computed using a bootstrap size of *ℬ* = 100. As a baseline estimate, we assume an overall California COVID-19 infection rate *π*_*I*_ of 2%, a rough estimate of the infection rate of the 1918 Influenza Pandemic [19] in the U.S., that is estimated to have had a basic reproductive number (*ℛ*_0_) comparable to that of COVID-19 [20, 21]. However, there is still substantial uncertainty surrounding the true COVID-19 infection rate primarily due to the lack of testing and the large prevalence of asymptomatic cases. Recent studies suggest that the true overall infection rate in the U.S. is much higher than what was initially hypothesized [22, 23].

Figure 2 depicts T-IR estimates and error bars indicating two bootstrap standard errors (SEs) for different combinations of gender and age group under the assumption of an overall California infection rate of 2%. The T-IR estimates range from 0.1% to 8.3% for female and 0.1 % to 8.9% for male. The figures present 6 different race/ethnicity groups: LatinX/ Hispanic (LatinX), White/ Caucasian (White), Asian, African American / Black (AA), Multi-Race, and American Indian or Alaska Native (AIAN). LatinX has the highest T-IRs, followed by African American. Both females and males age 80 and older have extremely high T-IRs compared with other age groups across race/ethnicity groups. T-IRs were non-monotonic at younger ages, with age groups 60–64 and 70–74 having slightly lower T-IRs than the preceding age groups, 50–59 and 65–69 respectively. Males had higher T-IRs than females across all age groups with the gender gap slightly increasing with age.

**Figure 2:**
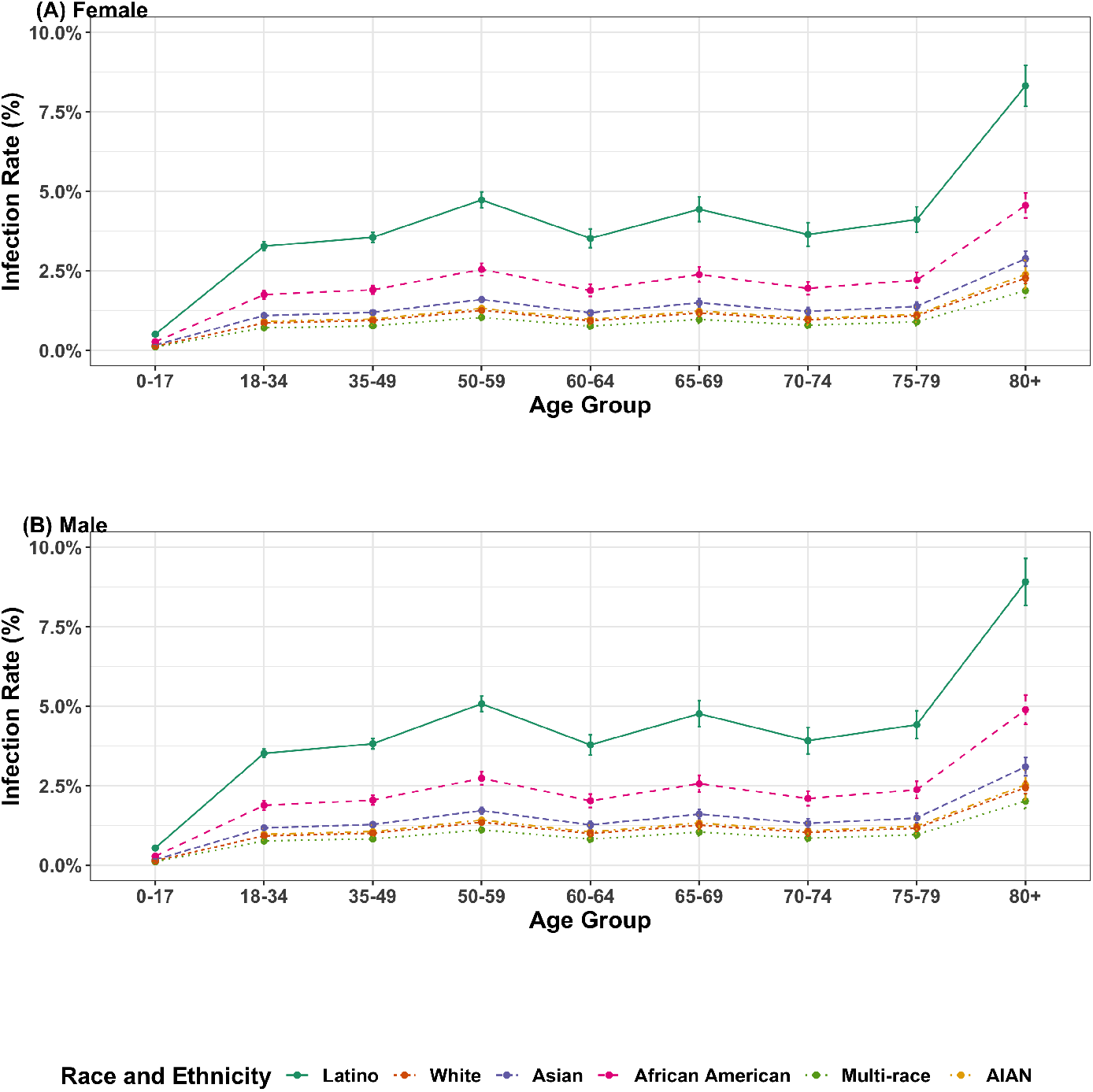
Estimated test-based infection rates given age and race/ethnicity, stratified by gender. (A) and (B) present the bootstrapped mean infection rates for female and male respectively. Only 6 racial and ethnicity groups are considered in the figures, including LatinX/ Hispanic (LatinX), White/ Caucasian (White), Asian, African American / Black (AA), MultiRace, and American Indian or Alaska Native (AIAN). The overall infection rate was assumed to be 2%, and the error bars denote two bootstrap standard errors.

We also considered alternate values for the overall California IR. Table 4 presents the point estimates and associated two SE intervals of the marginal T-IRs for gender and age group obtained from marginalization formula (20) assuming overall IRs of 1%, 2%, and 5%. The estimated marginal T-IRs for gender, age groups and race and ethnicity groups are consistent with the results presented in Figure 2, including males and older individuals having higher estimated T-IRs.

**Table 4:**
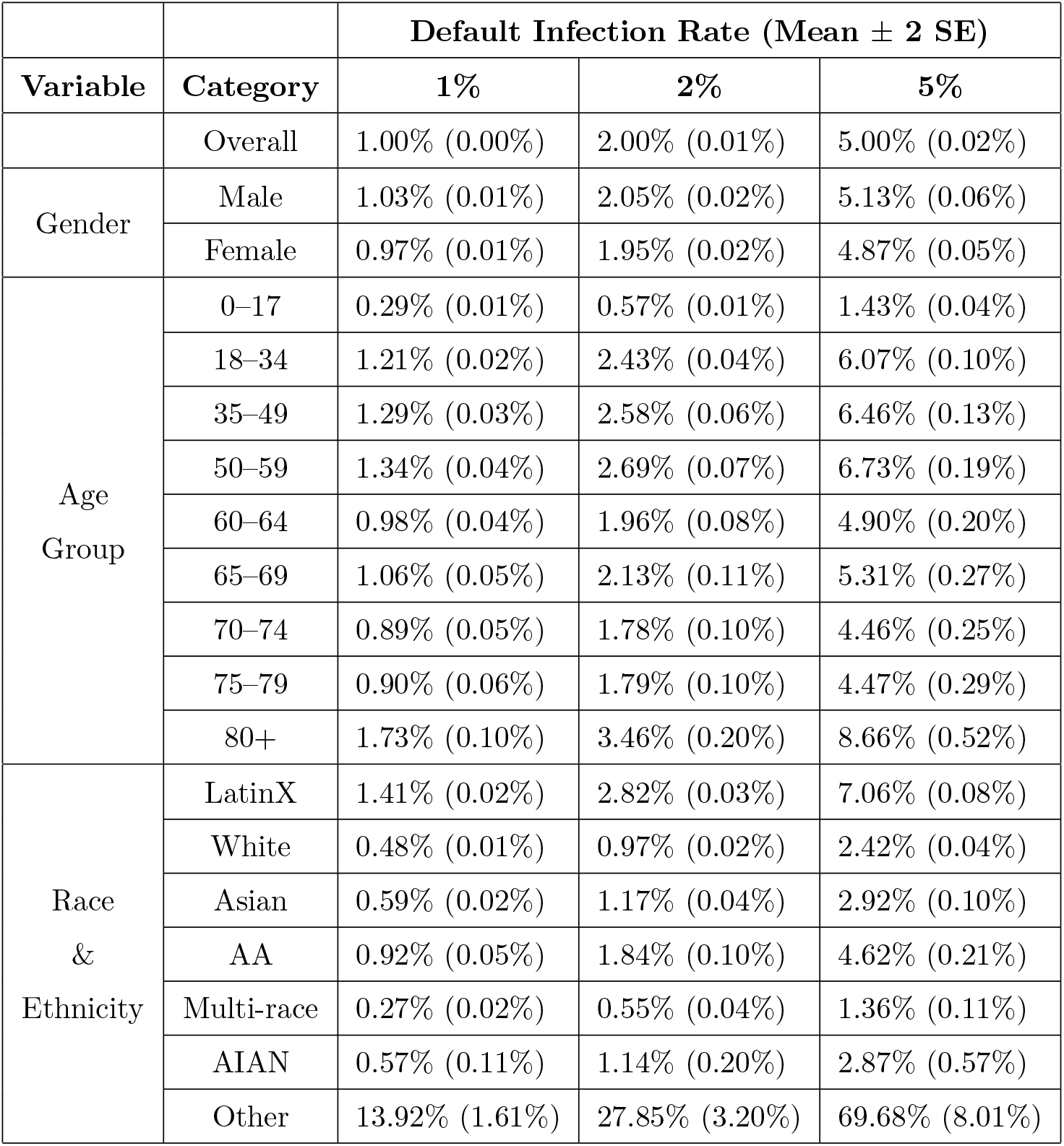
Estimated marginal test-based infection rates for different overall infection rates

Table 5 presents the point estimates and associated two SE intervals of the bootstrap estimated marginal T-CFRs obtained from marginalization formula (21), assuming an overall infection rate of *π*_*ℐ*_ = 2%; T-CFR estimates do not vary in expectation for different values of *π*_*ℐ*_. Males have a mean T-CFR 0.78% higher than females, and T-CFRs increase with age, ranging from less than 0.11% for the 0-34 age group to over 27.45% for the 80+ age group. Among 6 race and ethnicity groups, African American, Asian, and White are high-risk groups with mean T-CFRs as 7.42%, 6.80%, and 6.78% respectively. Other, LatinX, and Multi-race subgroups have T-CFRs below the overall 3.68% T-CFR for California.

**Table 5:** Estimated marginal test-based case fatality rates

| Variable | Category | Case Fatality Rate<br>(MEAN ± 2 SE) |
| --- | --- | --- |
| Overall | Overall | 3.68% (0.06%) |
| Gender | Male | 4.06% (0.13%) |
|  | Female | 3.28% (0.12%) |
| Age<br>Group | 0–34 | 0.11% (0.03%) |
|  | 35–49 | 0.74% (0.10%) |
|  | 50–59 | 2.07% (0.19%) |
|  | 60–64 | 4.08% (0.46%) |
|  | 65–69 | 7.41% (0.70%) |
|  | 70–74 | 11.25% (0.91%) |
|  | 75–79 | 17.19% (1.21%) |
|  | 80+ | 27.45% (0.87%) |
| Race<br>&<br>Ethnicity | LatinX | 2.62% (0.10%) |
|  | White | 6.78% (0.28%) |
|  | Asian | 6.80% (0.50%) |
|  | AA | 7.42% (0.71%) |
|  | Multi-race | 3.31% (1.05%) |
|  | AIAN | 5.19% (2.54%) |
|  | Other | 0.37% (0.10%) |

Figure 3 presents the estimated T-CFRs, obtained from formula (21), with error bars displaying two SEs of uncertainty for different combinations of gender and age groups, stratified by 6 race and ethnicity groups as shown in Figure 2. Males have higher estimated T-CFRs than females across all age-race levels, and the gender gap increases with age. African American and then Multi-race have higher estimated T-CFRs than other race groups in general across different age groups based on the stratified results. African American female even has a higher of T-CFRs than AIAN, LatinX and White male for each age group correspondingly.

**Figure 3:**
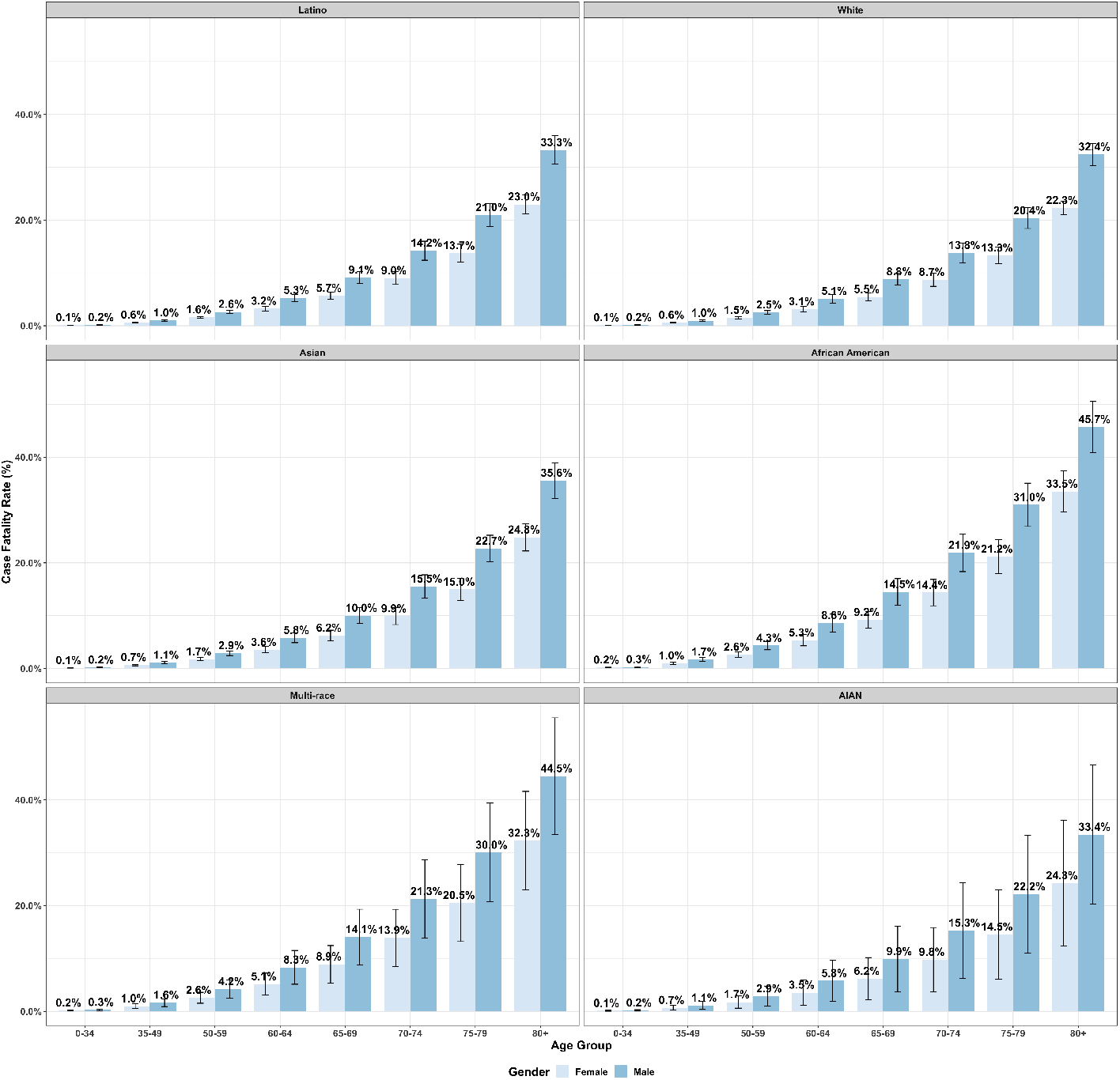
Estimated test-based case fatality rates by age and gender, stratified by race/ethnicity. The bootstrap mean case fatality rates are presented separately for LatinX, White, Asian, African American, Multi-Race, and AIAN groups. The overall infection rate was assumed to be 2%, and the error bars denote two bootstrap standard errors.

Although Multi-race has higher T-CFRs at each age groups than White, the overall marginal T-CFRs for Multi-race is lower than White as shown in Table 5. The reversal of the inequality between the size ratios is an example of Simpson’s paradox. The Multi-race population is younger than White population in California. For example, we can compare the number of adults (18 and above) and the number of children and adolescents (0 to 17 years old) within each race and ethnicity group. An overall adult-child ratio is defined as the total number of adults over that of children and adolescents ignoring race and ethnicity groups. Multi-race population in California has 0.43 times overall adult-child ratio (1.7% of adults and 4% of children and adolescents), while it is 1.3 times overall adult-child ratio for White population (38.8% of adults and 29.2% of children and adolescents)[6]. Since the T-CFRs increase with the age, a higher adult-child ratio in the age structure for Multi-race leads to a higher marginal T-CFR. The case still holds when we have multiple age groups. A similar paradox happens for LatinX and Asian. Meanwhile, the small proportions of Multi-race and AIAN in the general population also result in large error bars.

## 5. Discussion

In this paper, we combined aggregate COVID-19 case and fatality data with population demographic data in a pseudo-likelihood based multivariable logistic regression approach for obtaining early estimates of COVID-19 T-IRs and T-CFRs for subgroups of the California population. Overall, our results revealed that males, the elderly, and LatinX are marginally at relatively higher risk of COVID-19 infection, and that males, the elderly, Africa Americans, Asians, and Whites are marginally at elevated risk of mortality after COVID-19 infection. However, due to the imbalance in the age distribution of different races in California, the subgroups with the top 5 T-CFRs are Africa American male,

Multi-race male, Asian male, African American female, and AIAN male for each age group. Overall, therefore, African Americans are the race/ethnicity group most vulnerable to COVID-19 in California. We also found that the elevated infection and mortality risk for males and the greater mortality risk for all races increase with age.

The proposed methods are subject to three general limitations. First, the analysis is based on publicly available test-based infection rates and case fatality rates. It has been well documented that the lack of testing for COVID-19 in the U.S. has hindered efforts to estimate the true COVID-19 infection rate. Further compounding this issue is the high prevalence of asymptomatic COVID-19 cases. These two issues may lead to substantial underestimates of the infection rates and/or substantial overestimates of the case fatality rates from our analyses. Second, race/ethnicity is missing in 29% of the reported cases from CDPH, which may bias our estimates. Even though CDPH releases summary statistics for age-race covariates and our model can fit the finer data, we fit the marginal statistics for each risk factor in the analysis to minimize the impact of the potential sampling bias. Moreover, the case and fatality data released by CDPH provide marginal summary statistics for a subset of risk factors, and we do not have direct information on the joint distribution of all risk factors. Although the central goal of our proposed methods is to circumvent this limitation, the absence of direct multivariate information on the risk factors of COVID-19 infection and mortality as well as the sampling bias should be taken into account when interpreting the results of our models. Third, in this paper, we do not consider regularity conditions ensuring concavity associated with the pseudo-log-likelihood functions constructed in Equations (6) and (15), nor do we examine the asymptotic properties of the parameter estimates in Equations (7) and (16). Future research investigating the mathematical theory of the proposed methods is warranted.

Another promising avenue for future work is combining this method with a COVID-19 prediction model [24] to provide detailed demographic projections of COVID-19 cases and mortalities. This would be a substantial improvement over most COVID-19 prediction models, as they tend to be quite limited in their ability to forecast the demographic characteristics of the infected.

In summary, this paper provides a pragmatic tool for producing early estimates of COVID-19 T-IRs and T-CFRs for the California population, which offer valuable information to guide health policies concerning the control and prevention of COVID-19. In addition, our methods can be generalized into a general framework for early estimation of subpopulation IRs and CFRs from aggregate case and fatality data in other locations and for future epidemics.

## Data Availability

The data underlying the results presented in the study are based on the most recent COVID-19 case and fatality data from the California Department of Public Health (CDPH) and population-level demo-graphic data from the California Health Interview Survey (CHIS).

https://www.cdph.ca.gov/Programs/CID/DCDC/Pages/COVID-19/Race-Ethnicity.aspx

https://update.covid19.ca.gov/#top

https://www.cdph.ca.gov/Programs/CID/DCDC/Pages/COVID-19/COVID-19-Cases-by-Age-Group.aspx

https://healthpolicy.ucla.edu/chis/data/Pages/GetCHISData.aspx

## Acknowledgements

We thank Dr. Sudipto Banerjee and Jay J. Xu (University of California, Los Angeles) for their many helpful comments and assistance.

## Author Contributions

DX, MAS, and CMR conceptualized the study design. DX, LZ, GLW, and CMR drafted the original manuscript. DX, PS, TB, and CMR performed the data collection and literature search. DX and LZ developed the methodology, conducted the statistical analysis, and interpreted the results. DX, LZ, GLW, PS, TB, JZ, JS, and MAS performed the computer programming. All authors revised the manuscript and approved the final version.

## Funding

The authors received no specific funding for this work.

## Declaration of Competing Interest

The authors have declared that no competing interests exist.

## Notes

### Competing Interest Statement

The authors have declared no competing interest.

### Author Declarations

No IRB body is required for this study.

